# DNA Methylation Age Acceleration Mediates the Relationship between Systemic Inflammation and Cognitive Impairment

**DOI:** 10.1101/2024.07.24.24310948

**Authors:** César Higgins Tejera, Peiyao Zhu, Erin B. Ware, Margaret T. Hicken, Matthew Zawistowski, Lindsay C. Kobayashi, Dominika Seblova, Jennifer Manly, Bhramar Mukherjee, Kelly M. Bakulski

## Abstract

**Background:** Chronic inflammation and DNA methylation are potential mechanisms in dementia etiology. The linkage between inflammation and DNA methylation age acceleration in shaping dementia risk is understudied. We explored the association of inflammatory cytokines with cognitive impairment and whether DNA methylation age acceleration mediates this relationship.

**Methods:** In a subset of the 2016 wave of the Health and Retirement Study (n=3,346, age>50), we employed logistic regression to estimate the associations between each inflammatory cytokine (interleukin-6 (IL-6), C-reactive protein (CRP), and insulin-like growth factor-1 (IGF-1)), and both Langa-Weir classified cognitive impairment non-dementia and dementia, respectively. We calculated DNA methylation age acceleration residuals by regressing GrimAge on chronologic age. We tested if DNA methylation age acceleration mediated the relationship between systemic inflammation and cognitive impairment, adjusting for sociodemographic, behavioral factors, chronic conditions, and cell type proportions.

**Results:** The prevalence of cognitive impairment was 16%. In the fully-adjusted model, participants with a doubling of IL-6 levels had 1.12 (95% CI: 1.02-1.22) times higher odds of cognitive impairment. Similar associations were found for CRP and IGF-1. Participants with a doubling of IL-6 levels had 0.77 (95% CI: 0.64, 0.90) years of GrimAge acceleration. In mediation analyses with each cytokine as predictor separately, 17.7% (95% CI: 7.0%, 50.9%) of the effect of IL-6 on cognitive impairment was mediated through DNA methylation age acceleration. Comparable mediated estimates were found for CRP and IGF-1.

**Conclusions:** Systemic inflammation is associated with cognitive impairment, with suggestive evidence that this relationship is partially mediated through DNA methylation age acceleration.

## Introduction

The risk of cognitive impairment in aging adults is influenced by various health determinants (1–3). Peripheral immunity may be related to cognitive impairment in older adults (4,5). Molecular mechanisms, such as DNA methylation age acceleration, may contribute to poor cognitive health outcomes, including cognitive impairment (6). Therefore, unveiling the potential shared contributions of the immune response and the cellular mechanisms associated with accelerated epigenetic aging could inform public health efforts to identify individuals at greater risk for cognitive decline.

The immune profiles of aging adults are characterized by a state of immune dysregulation with high circulating blood levels of pro-inflammatory cytokines in the absence of tissue triggers - a process often referred to as *inflammaging* (7). Elevated levels of chronic inflammation are associated with multiple age-related diseases such as coronary heart disease, stroke, diabetes, kidney disease, and dementia (2,8–11). High levels of pro-inflammatory cytokines such as C-reactive protein (CRP), Interleukin-6 (IL-6), Interleukin-1 (IL-1), and Tumoral Necrosis Factor alpha (TNF-α) have been found in atherosclerotic plaques of coronary arteries, tubular epithelial cells of the kidney tissue, and in brain parenchyma post ischemic stroke (7). The co-localization of pro-inflammatory cytokines in tissue-specific processes suggests that systemic inflammation is an underlying mechanism of multiple chronic diseases and a biological signature of accelerating aging. However, whether inflammaging is a contributor to aging-related pathologies, such as those leading to cognitive impairment, remains uncertain.

To better understand the role of systemic inflammation in the etiology of age-related diseases, we need to assess the link between the inflammatory response and molecular mechanisms implicated in accelerated aging (7,12,13). DNA methylation clocks are biomarkers of biological aging (14). These clocks are built from machine learning algorithms that identify patterns of specific DNA methylation sites in different tissues, which are then used to predict the DNA methylation age of the sample source (15,16), The residual from a regression of an individual’s estimated DNA methylation age and their chronological age is a measure of accelerated biological aging. The first generation of DNA methylation clocks were designed to estimate chronologic age and the second generation of DNA methylation clocks sought to characterize phenotypic age, which is of primary interest to this study. Premature biological aging, as measured by the second generation DNA methylation clocks, has been associated with several chronic conditions, early mortality, and cognitive decline (15,17,18). To date, epidemiological studies exploring the interplay between blood circulating levels of cytokines, DNA methylation clocks, and aging-related diseases are limited.

In a cross-sectional sample of the Health and Retirement Study, a population-based cohort of older United States adults, we tested the hypothesis that elevated circulating blood levels of cytokines and accelerated DNA methylation aging would be associated with prevalent cognitive impairment. We used mediation analysis to estimate to what extent the effect of each cytokine on cognitive impairment was mediated through DNA methylation age acceleration.

## Methods

### Study population

Data were from the US Health and Retirement Study (HRS), a population-based longitudinal study of aging adults sponsored by the National Institute on Aging (NIA U01AG009740). To ensure a representative sample of the demographic composition of the US, the HRS oversamples non-Hispanic Black and Hispanic participants employing a multi-stage probability design (19). The original HRS cohort began in 1992, and participants are interviewed every two years. The study interview collects data on sociodemographic, economic, behavioral, psychological, and other health-related factors thought to be associated with the aging process (20). In 2016, all participants who completed the interview were asked to consent to a Venous Blood Study, except for proxy participants and those living in a nursing home. A panel of seven cytokines and other markers of cellular aging were assessed in subsamples of the Venous Blood Study (21,22). Participants provided written informed consent at the time of the data collection. These secondary data analyses were approved by the University of Michigan Institutional Review Board (HUM00128220).

In a subsample of the 2016 Venous Blood Study, we tested the cross-sectional associations between seven cytokines (exposures) and six DNA methylation clocks (mediators); and their respective associations with prevalent cognitive impairment (outcome). In total, 9,934 participants from HRS study were selected for the venous blood study in 2016, and 3,556 participants had complete information on cytokines, DNA methylation clocks and APOE-*ε4* allele status. Among these, 3,450 had complete demographic information. An additional 101 participants were excluded due to self-reporting their race/ethnicity as “non-Hispanic Other”. The final sample included 3,346 participants, and participant inclusion is visualized in a flow chart (**Supplemental Figure 1**). This study adhered to both the Strengthening the Reporting of Observational Studies in Epidemiology (STROBE) guidelines (23) and the Guideline for Reporting Mediation Analyses (AGReMA) (24).

### Cytokine assays

Cytokine assays were conducted on the Venous Blood Study sample of the 2016 HRS wave. Blood collection was managed by Hooper Holmes Health & Wellness. Approximately 50.5mL of blood were collected in 6 tubes – one 8 mL CPT tube, three 10 mL double gel serum separator tubes (SST), one 10 mL EDTA whole blood tube, and a 2.5 mL PAXgene RNA tube. The SST tubes were centrifuged in the field before being shipped overnight to the Advance Research Diagnostic Laboratory at the University of Minnesota. Tube processing was done within 24 hours of arrival at the lab or within 48 hours of collection. A panel of seven cytokines (CRP, IL-6, IL-1RA, IL-10, sTNF-R1, IGF-1, and TGF-β1) were measured in serum using enzyme-linked immunosorbent assay (ELISA). Detailed information on each cytokine assay procedure can be found elsewhere (21,22).

In statistical analyses, we used cytokines as continuous variables. Given their positive skewness and the wide range of cytokine levels across participants, we used the log2 transformation of CRP, IL-6, IL-1RA, IL-10, sTNF-R, and the square root of IGF-1 to normalize their distribution. We considered patterns among proinflammatory cytokines (CRP, IL-6, IL-10, and sTNF-R1) and among anti-inflammatory cytokines (IGF-1 and TGF-β1).

### DNA methylation assays and DNA methylation age acceleration measures

DNA methylation was collected and processed by the HRS (21,22). DNA methylation was measured in a non-random subsample (n=4,104) of participants who consented to participate in the Venous Blood Study, and a total of 4,018 (97.9%) samples passed quality control assessment. This subsample was representative of the sociodemographic distribution of the HRS. Whole genome DNA methylation data were assessed using the 850K MethylationEPIC BeadChip (Infinium) microarray (Illumina Inc.) (21) at the University of Minnesota. Samples were randomized across plates by key demographic variables such as age, cohort, sex, education, and race/ethnicity. The R package *minfi* (21,25) was used to preprocess the RGChannelSet object and perform quality control. Beta methylation values were calculated. Sample mismatches in sex and any control (cell lines, blind duplicates) were removed (n=58). For any given participant, probes with detection p-values <0.01 were imputed to the mean methylation value of the given probe across all samples (21,22).

The HRS calculated and released data for several epigenetic clocks. A DNA methylation clock is an estimator built from DNA methylation CpG sites that are strongly correlated with age (15). DNA methylation clocks are generally categorized as: a) chronological, or clocks that provide a “forensic” estimation of chronological age (i.e., Horvath, Hannum, HorvathSkin); and b) phenotypical, or clocks that are informative of a disease state (i.e., GrimAge, Levine, and the methylation pace of aging clock or DunedinPoAm) (15). For the purpose of our analysis, we used these six clocks and explored their relationship with both inflammatory cytokines and cognitive impairment. In our mediation analysis, we prioritized the phenotypic clock GrimAge for its strong *a priori* relationship with both cytokines and cognitive impairment (26). The GrimAge clock consists of approximately 1,030 CpGs and was built using smoking pack-years and seven plasma proteins associated with mortality risk (27). Horvath is a multi-tissue clock consisting of 353 CpGs and an average error of 3-5 years (16,18). The Hannum clock reflects age-related changes in cell compositions and consist of 71 CpGs (28). Despite sharing only 6 CpG sites, Horvath and Hannum clocks are highly predictive of all-cause mortality (17). The skin and blood clock (HorvathSkin) was built from 391 CpGs, and shares 71 CpGs with the Hannum and 60 CpGs with the Horvath clock, this clock was developed to measure the age of human fibroblasts, keratinocytes, buccal cells, endothelial cells, skin and blood samples (29). The Levine clock, also known as PhenoAge, was built from 513 CpGs that predict a composite score of biological aging based on diverse clinical parameters (30). Finally, the age-38 DunedinPoAm estimator was built to predict longitudinal changes in 18 physiological markers of organ function (31). We transformed the DunedinPoAm rate into years for alignment with the other age estimators.

DNA methylation age acceleration represents the residuals of the relationship between estimated DNA methylation age and observed chronological age (6). This variation reflects an individual’s rate of aging (17). To estimate age accelerated residuals, we regressed each DNA methylation clock (Y) on participants’ chronological age (X) at the HRS 2016 wave. DNA methylation age accelerated residuals were inspected to meet linear regression assumptions (e.g. normality and constant variance of the errors). Predicted residual values greater than zero reflect DNA methylation age acceleration and negative or zero residual values reflect non-accelerated DNA methylation age. In statistical analysis, we used epigenetic age accelerated residuals in their continuous form.

### Cognitive Status

All participants in our study were non-proxy respondents. Cognitive function was assessed through the a series of telephone interview cognition questions (32), which provides a total score of 0 to 27 points based on a battery of cognitive test that include a 10-word immediate and delayed recall test, a serial 7 subtraction, and counting backwards. Participants’ cognitive status was categorized using the Langa-Weir algorithm, which uses a 27-point scale derived from cognition-related questions. The classification were dementia (0 to 6 points), cognitive impairment non-dementia (7 to 11 points), and normal cognition (12 to 27 points) (33). In statistical analyses, we compared dementia and cognitive impairment non-dementia versus normal cognition separately. To leverage a larger number of cases and higher statistical power to detect potentially mediated effects in mediation analysis, we combined dementia and cognitive impairment non-dementia into a single category representing cognitive impairment and compared it to normal cognition.

### Covariate measures

Our analysis adjusted for potential confounders in the association between inflammatory biomarkers and cognitive impairment, and between DNA methylation age acceleration and cognitive impairment. Sociodemographic and genetic confounders included: age at interview date (continuous), self-reported sex (female, male), self-reported racialized social group (Hispanic, non-Hispanic Black, non-Hispanic White), self-reported highest educational attainment (more than college, college or some college, high school or less), and *APOE*-*ε4* allele carrier status (at least 1 copy, no copy). Genetic information on *APOE*-*ε4* allele status was obtained from phased genetic data imputed to the worldwide 1000 Genomes Project reference panel. Genotyping and imputation information for the HRS is available elsewhere (34). Health behavioral confounders included: self-reported body mass index (BMI; calculated as weight kilograms divided by height in meters squared, continuous), weekly vigorous, moderate, or light physical activity (yes, no), alcohol consumption (reported as number of drinks a day when drinks, with non-drinkers assigned a value of 0, continuous), smoking status (current, former, never). Chronic health conditions included self-reports of a physician diagnosis of each of the following conditions: high blood pressure (yes, no), diabetes (yes, no), cancer (yes, no), lung disease (yes, no), heart disease (yes, no), stroke (yes, no), psychiatric problems (yes, no), and arthritis (yes, no); operationalized in our models as a categorical variable (0, 1-2, 3 or more conditions). For the relationship between inflammatory biomarkers and DNA methylation age acceleration, we adjusted for age, sex, racialized social group, highest educational attainment as mentioned above. Additionally, we adjusted for cell type proportions (percent granulocytes and percent lymphocytes measured from complete blood count measures, both continuous variables). In the mediation analysis, we adjusted for all these covariates in our model.

### Statistical Analysis

We compared the covariate distributions between included and excluded HRS participants due to missing data. First, in bivariate analysis, we examined the distribution of covariates by cognitive status (dementia, cognitive impairment non-dementia, and normal cognition) using one-way ANOVA test for continuous variables and chi-square test for categorical variables. We estimated the Pearson correlation between cytokines, epigenetic clocks, and cell type proportions and visualized them in correlation plots.

### Cytokine and cognitive status regression models

We employed multivariable-adjusted logistic regression analysis to estimate the association between each cytokine exposure and the odds of each of prevalent dementia and cognitive impairment non-dementia relative to normal cognition. A fully adjusted logistic regression model was built for each combination of cytokine and cognitive status. This model included adjustments for sociodemographic factors (age, sex, racialized social group, education), *APOE-ε4* carrier status, health behaviors (body mass index, smoking status, alcohol consumption, exercise), and chronic conditions. We reported the odds ratio (OR) and 95% confidence interval (CI) for each cytokine. To account for multiple comparisons, we calculated false discovery rates (FDR) (35), considering an FDR value of less than 0.1 as significant.

### Cytokines and DNA methylation age acceleration models

We employed multivariable linear regression analysis to estimate the associations between each cytokine and each DNA methylation age acceleration measure. We built three generalized linear models to estimate the relationship between each cytokine with each phenotypic clock (GrimAge, Levine, DunedinPoAm) and each chronologic clock (Horvath, Hannum, HorvathSkin). We reported the effect estimates and 95% CI for each cytokine and calculated the FDR.

### DNA methylation age acceleration and cognitive status models

We used multivariable logistic regression analyses to estimate the associations between each DNA methylation clock and cognitive status. In this analysis, we adjusted for sociodemographic factors (age, sex, racialized social group, education), *APOE-ε4* carrier status, cell type proportions, health behaviors (body mass index, smoking status, alcohol consumption, exercise), and chronic conditions. We reported the OR and 95% CI for cognitive impairment associated with a 1-year increase of DNA methylation age acceleration and calculated the FDR.

### Mediation analysis

We employed regression-based causal mediation analysis (36,37) to investigate whether DNA methylation age acceleration mediated the total effect of cytokine levels on cognitive impairment (dementia and cognitive impairment non-dementia) relative to normal cognition.(38) For our mediation models, we selected three different cytokines (CRP, IL-6, and IGF-1) as exposures, because these biomarkers were strongly associated in the multivariable models with both dementia and cognitive impairment non-dementia. We also selected GrimAge acceleration as a mediator because this clock was the most strongly associated in multivariable logistic regression models with impaired cognitive status. To leverage a larger sample size and statistical power, we combined dementia and cognitive impairment non-dementia cases into a single category representing cognitive impairment, our outcome.

We built three different mediation models with C-reactive protein, interleukin-6, and IGF-1 as the primary predictor separately. We used the *CMAverse* R studio package to estimate the proportion of the total effect of each cytokine on cognitive impairment that was mediated through the GrimAge clock (36,37). The *CMAverse* accommodates for different generalized linear models within the mediation analysis. For example, logistic regression was employed to estimate the exposure-outcome relationship. The exposure-mediator relationship was estimated through linear regression analysis, and the mediator-outcome relationship was estimated through logistic regression.

In addition, because our primary interest in this mediation analysis was to estimate the mediated effect of epigenetic age on the relationship between cytokine exposure and cognitive impairment, we calculated mediational e-values to understand to what extent an unmeasured confounder could nullified the observed mediational proportions (39). All statistical analyses were conducted using R statistical software (version 3.6.2). Code to perform the analyses is available (https://github.com/bakulskilab).

## Results

### Sample characteristics

Our study included a cross-sectional sample of 3,346 participants of the 2016 venous blood study in the HRS cohort (**Supplemental Figure 1**). The included sample participants were older and had a relatively higher prevalence of chronic conditions compared to the excluded sample, while other characteristics were generally similar across both groups (**Supplemental Table 1**). On average, included participants were 70 (SD=9.6) years of age, 58% female, 70% had completed high school education or less, and 17% of participants were racialized as non-Hispanic Black and another 14% as Hispanic (**Table 1**). The prevalence of dementia was 3.7% (n=125) and the prevalence of cognitive impairment non-dementia was 16% (n=543). Elevated levels of pro-inflammatory cytokines such as CRP, IL-6, IL-10, and sTNF-R1 were observed among cognitively impaired participants, but the differences were not statistically significant (**Table 1 & Figure 1**). In contrast, higher levels of anti-inflammatory cytokines such as IGF-1 and TGF-β were observed in cognitively normal participants. There were no differences in the distributions of IL-1ra across the different cognitive categories (**Table 1**). Higher GrimAge acceleration was observed among participants with dementia (1.0 years) and cognitive impairment non-dementia (1.13 years), relative to those with normal cognition (-0.27 years) (**Table 1**).

**Figure 1:**
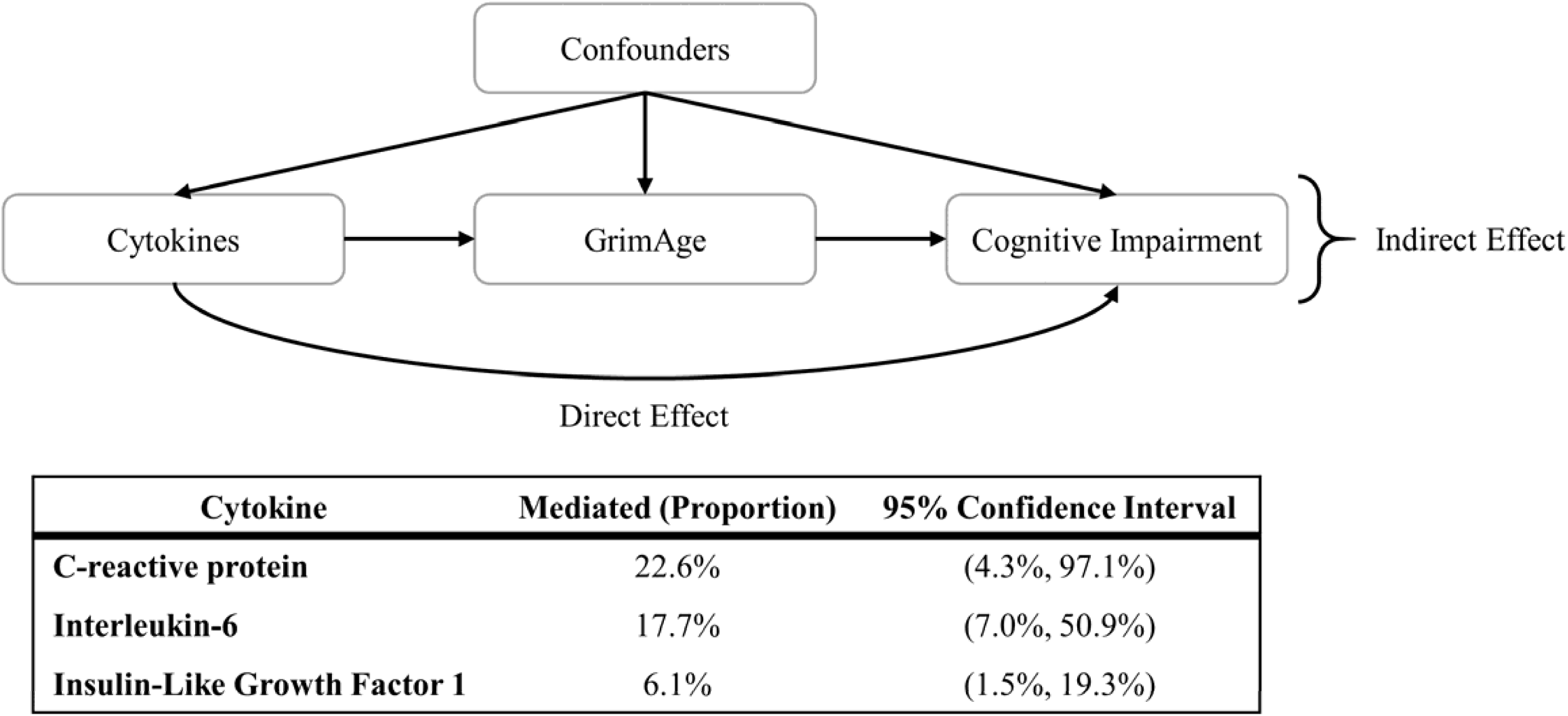
Directed Acyclic Graph illustrating the hypothesized relationship between cytokines, GrimAge acceleration, and cognitive impairment. Mediation estimates represent the proportion of the total effect of a cytokine on cognitive impairment that is due to DNA methylation age acceleration. Confounders include socio- demographic variables (age, sex, racialized social group, education), behavioral factors (body mass index, exercise, smoking status, alcohol consumption), chronic conditions, APOE- ε4 allele carrier status, and cell type proportions (percent granulocytes and lymphocytes) in the Health and Retirement Study 2016 wave (n=3,346).

**Table 1:**
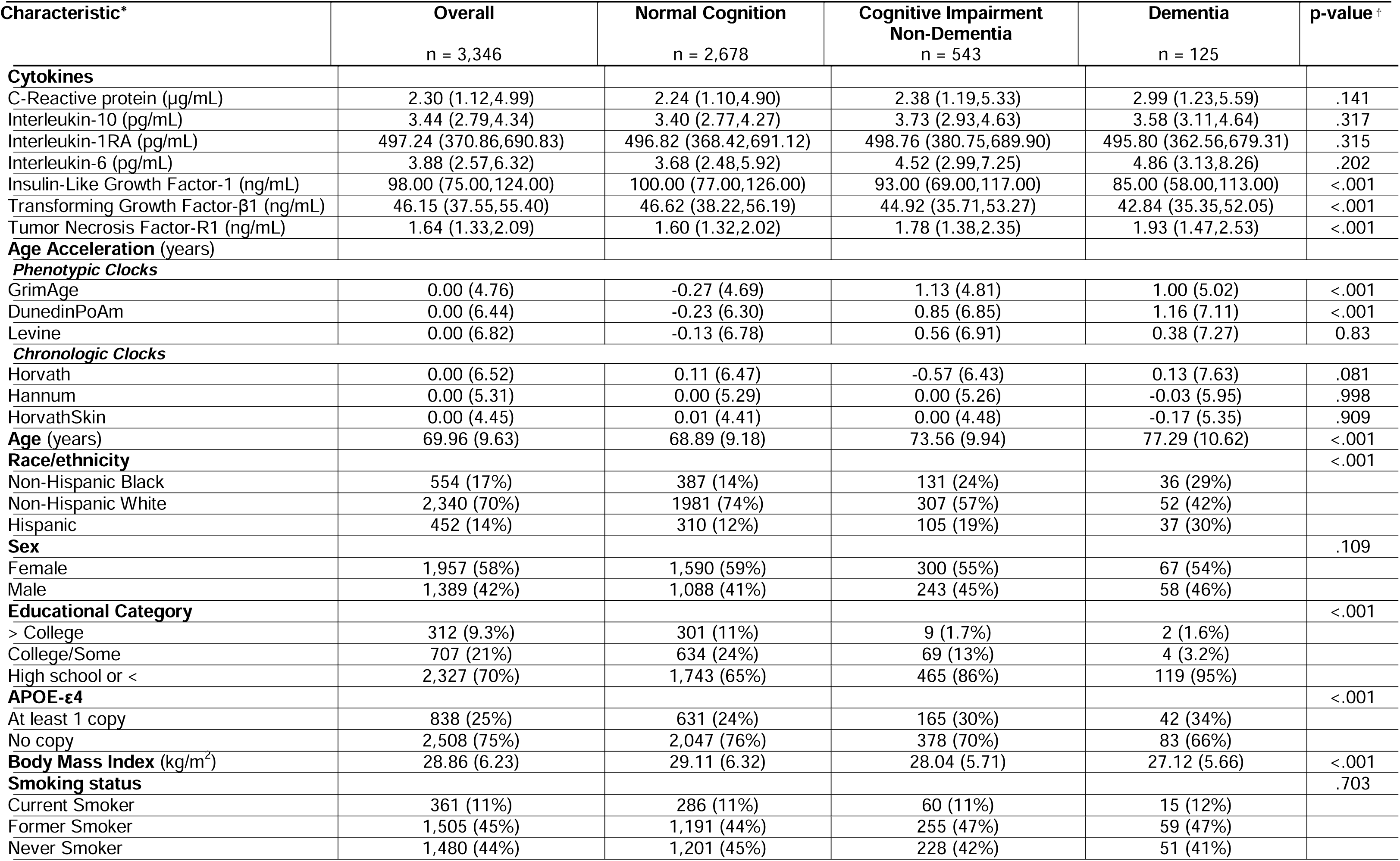
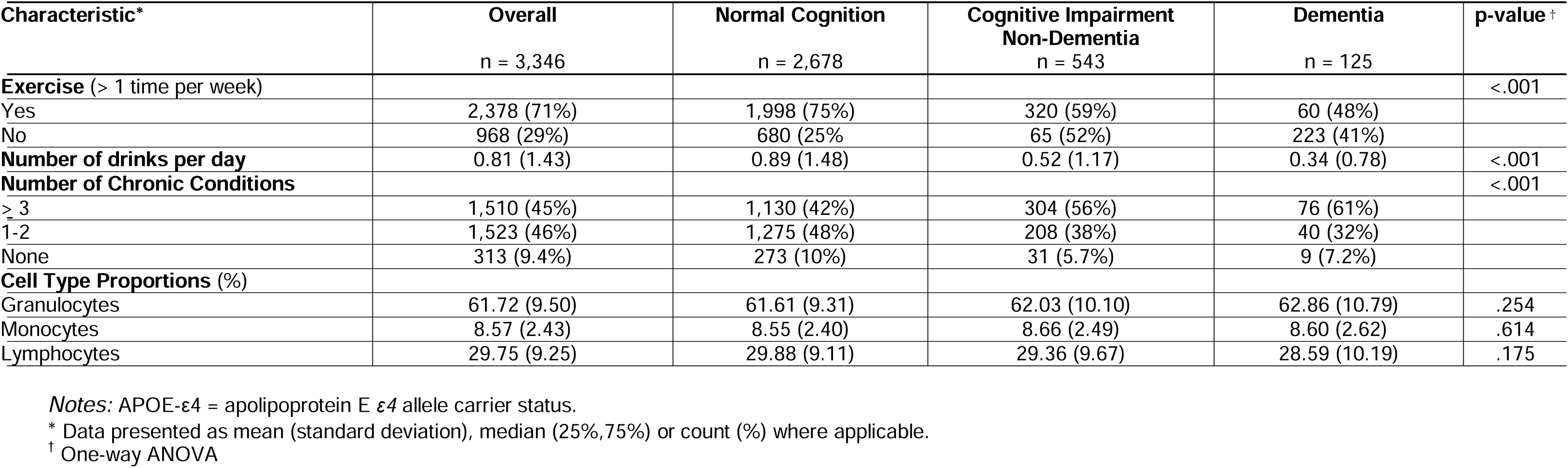
Sample distribution of cytokines, DNA methylation age acceleration, sociodemographic, behavioral, chronic conditions, and genetic characteristics and cell type proportions by cognitive status in the Health and Retirement Study, wave 2016 by cognitive status

Proinflammatory cytokines (i.e., CRP, IL-6, IL-10, and sTNF-R1) were weakly to moderately correlated (**Supplemental Figure 2**). Among all cytokines, CRP and IL-6 exhibited the strongest correlation (r = 0.44). The anti-inflammatory cytokine IGF-1 was negatively correlated with pro-inflammatory cytokines (i.e., IL-10, IL1-ra, sTNF-R1, CRP, IL-6) (**Supplemental Figure 2**). The phenotypical clocks GrimAge and DunedinPoAm had the strongest correlation coefficient (r = 0.62) and the chronological clocks such as Horvath, Hannum, and HorvathSkin were moderately correlated (**Supplemental Figure 2**).

### Adjusted association between cytokines and cognitive status

In general, proinflammatory cytokines were associated with higher odds of prevalent dementia and cognitive impairment non-dementia. For example, a doubling of CRP was associated with a 1.16 (95% CI: 1.01, 1.32) times greater odds of prevalent dementia with a statistically significant FDR adjusted P-value, and 1.07 (95% CI: 1.00, 1.15) times higher odds of cognitive impairment non-dementia (**Table 2**). Similarly, a doubling of IL-6 was associated with 1.19 (95% CI: 1.00, 1.41) times greater odds of prevalent dementia and 1.12 (95% CI: 1.02, 1.22) times greater odds of cognitive impairment non-dementia with significant FDR adjusted P-value. In contrast, the anti-inflammatory cytokine IGF-1 was associated with lower odds of prevalent dementia (OR = 0.88; 95% CI: 0.78, 0.98) with a statistically significant FDR adjusted P-value and cognitive impairment non-dementia (OR = 0.94; 95% CI: 0.89, 1.00). Other cytokines were not associated with cognitive status (**Table 2**).

**Table 2:** Prevalent Odds Ratios (ORs) from logistic regression analysis, estimates represent the association for each doubling of cytokine levels with prevalent dementia and cognitive impairment non-dementia, relative to normal cognition in the US Health and Retirement Study (2016).

| Cytokines | Dementia versus normal cognition<br>(n=2,803) |  |  | Cognitive impairment non-dementia<br>versus normal cognition (n=3,221) |  |  |
| --- | --- | --- | --- | --- | --- | --- |
|  | OR | [95%CI] | FDR | OR | [95%CI] | FDR |
| **C-Reactive protein\*** | 1.16 | [1.01,1.32] | 0.093 | 1.07 | [1.00,1.15] | 0.107 |
| **Interleukin-6\*** | 1.19 | [1.00,1.41] | 0.093 | 1.12 | [1.02,1.22] | 0.084 |
| **Interleukin-10\*** | 1.06 | [0.78,1.42] | 0.8 | 1.04 | [0.88,1.21] | 0.8 |
| **Interleukin-1RA\*** | 1.08 | [0.79,1.46] | 0.8 | 0.98 | [0.84,1.14] | 0.8 |
| **Insulin-Like Growth Factor-1<sup>†</sup>** | 0.88 | [0.78,0.98] | 0.093 | 0.94 | [0.89,1.00] | 0.107 |
| **Transforming Growth Factor-β1** | 1.00 | [0.98,1.01] | 0.8 | 1.00 | [0.99,1.00] | 0.42 |
| **Tumor Necrosis Factor-R1\*** | 1.10 | [0.77,1.52] | 0.8 | 1.12 | [0.94,1.33] | 0.35 |

Notes: OR = Odds Ratio; CI = confidence interval in brackets; FDR = false discovery rate. Model was adjusted for race, age, sex, education categories, APOE- ε4 allele status, smoking status, alcohol consumption, body mass index, exercise, and chronic conditions.
\* Variable log base 2 transformed.
<sup>†</sup> Squared root transformed.

### Association between cytokines and DNA methylation age acceleration

In multivariable adjusted models, cytokines were highly associated with DNA methylation age acceleration, and pro-inflammatory cytokines were correlated with increased age accelerated residuals (**Supplemental Table 2**). Exposure to anti-inflammatory cytokines was associated with decreased DNA methylation age acceleration. In our sample, a doubling of CRP was associated with 0.69 (95% CI: 0.54, 0.85) years of Levine age acceleration, 0.56 (95% CI: 0.47, 0.66) years of GrimAge age acceleration, and 0.77 (95% CI: 0.64, 0.90) years of DunedinPoAm age acceleration, all with significant FDR adjusted P values. Similar patterns of association were observed between CRP and chronologic age clocks (Horvath, Hannum, and HorvathSkin) (**Supplemental Figure 3** & **Supplemental Table 2**). Interleukin-6, IL-1RA, IL-10, and sTNF-R1 were also strongly associated with higher DNA methylation age acceleration across chronological and phenotypical clocks (**Supplemental Figure 3 & Supplemental Table 2**). In contrast, higher levels of the anti-inflammatory IGF-1 were associated with lower age acceleration across all phenotypic, but not the chronologic clocks. For example, a one unit increase in the square root-transformed IGF-1 levels was associated with a -0.19 (95% CI: -0.32, -0.07) lower years of Levine age acceleration, -0.14 (95% CI: -0.22, -0.06) lower years of GrimAge acceleration, and -0.32 (95% CI: -0.42, -0.21) lower years of DunedinPoAm acceleration (**Supplemental Figure 3 & Supplemental Table 2**). TGF-β was associated with DudeninPoAm clock. A doubling of TGF-β was associated with 0.02 (95%CI: 0, 0.03) years of DunedinPoAm age acceleration.

### Association between DNA methylation age acceleration and Cognitive Status

In multivariable logistic regression models, phenotypic clocks were associated with dementia and cognitive impairment non-dementia, but not chronological clocks. A one-year increase in GrimAge acceleration was associated, though not FDR significant, with 1.01 (95% CI: 0.96, 1.07) times higher odds of prevalent dementia and separately 1.05 (95% CI: 1.02, 1.08, FDR adjusted p-value=0.009) times higher odds of prevalent cognitive impairment non-dementia. An association between the DunedinPoAm clock and both dementia and cognitive impairment non-dementia was detected but became non-significant when fully adjusting for covariates (**Table 3**).

**Table 3:** Prevalent Odds Ratios (ORs) from logistic regression analysis, estimates represent the association between DNA methylation age accelerated and prevalent dementia and cognitive impairment non-dementia in the US Health and Retirement Study (2016)

| DNA methylation age acceleration (years) | Dementia versus normal cognition<br>(n=2,803) |  |  | Cognitive impairment non-dementia<br>versus normal cognition<br>(n=3,221) |  |  |
| --- | --- | --- | --- | --- | --- | --- |
|  | OR | [95%CI] | FDR | OR | [95%CI] | FDR |
| **Phenotypic Clocks** |  |  |  |  |  |  |
| GrimAge | 1.01 | 0.96, 1.07 | 0.932 | 1.05 | 1.02, 1.08 | 0.002 |
| DunedinPoAm | 1.01 | 0.97, 1.04 | 0.932 | 1.01 | 0.99, 1.03 | 0.45 |
| Levine | 1.00 | 0.97, 1.03 | 0.932 | 1.01 | 0.99, 1.02 | 0.4 |
| **Chronologic Clocks** |  |  |  |  |  |  |
| Horvath | 1.01 | 0.98, 1.04 | 0.932 | 0.99 | 0.97, 1.00 | 0.204 |
| Hannum | 1.02 | 0.98, 1.05 | 0.932 | 1.00 | 0.99, 1.02 | 0.72 |
| HorvathSkin | 1.00 | 0.96, 1.04 | 0.932 | 1.00 | 0.98, 1.02 | 0.9 |

*Notes:* OR = Odds Ratio; CI = confidence interval in brackets; FDR = false discovery rate. Model was adjusted for race, age, sex, education categories, APOE-ε4 allele status, cell components, smoking status, alcohol consumption, body mass index, exercise, and chronic condition.

### GrimAge Age Acceleration Mediates the Effect of Cytokines on Cognitive Impairment

In fully adjusted mediation models, we found that GrimAge acceleration mediated the effects of three different cytokines on cognitive impairment (dementia and cognitive impairment non-dementia combined). In a model with CRP as the primary exposure, GrimAge acceleration mediated 22.6% (95% CI: 4.3%, 97.1%) of the total effect of CRP on cognitive impairment (**Figure 1** & **Supplemental Table 3**). A different model with IL-6 as the primary exposure showed that GrimAge acceleration mediated 17.7% (95% CI: 7.0%, 50.9%) of the total effect of IL-6 on cognitive impairment. Lastly, a mediation model with IGF-1 as the primary exposure demonstrated that 6.1% (95% CI: 1.5%, 19.3%) of the total effect was mediated through GrimAge deacceleration (**Figure 1** & **Supplemental Table 3**). Our mediational e-values suggested that an unmeasured confounder associated with both CRP and cognitive impairment of an approximate magnitude of 1.14 could completely explain away the observed mediated effect, but a weaker confounder could not (**Supplemental Table 3**). To provide further context, observed confounders in our direct effect model such as age (OR: 1.07, 95% CI:1.05,1.08), race (Non-Hispanic Black, OR: 2.93, 95% CI = 2.25, 3.81; Hispanic, OR: 3.41, 95% CI = 2.61, 4.46), and education (High school or less, OR = 7.07, 95% CI = 3.94 to 14.1) (**Supplemental Table 4**) had larger odds ratios compared to the mediational E-value of 1.14. This indicated that the observed mediation effect is relatively weak to potential unmeasured confounders. Mediational e-values of similar magnitude were observed for IL-6, and IGF-1 (**Supplemental Table 3**).

## Discussion

In a large and diverse sample of older adults in the United States, we observed that higher circulating blood levels of pro-inflammatory cytokines and lower levels of anti-inflammatory cytokines were associated with cognitive impairment, and that these associations were partially mediated by accelerated biological aging (6.1% - 22.6% mediation), as assessed using DNA methylation clocks. These findings suggest a connection between peripheral systemic inflammation in neurodegenerative disorders such as Alzheimer’s disease and other dementias (40). Altogether, these findings suggest that accelerated DNA methylation aging may be an important cellular mechanism through which cytokines influence cognitive decline during aging.

Our results are consistent with other observational studies describing the association between blood circulating cytokine levels and cognitive impairment (41–45). In our sample, we found that higher blood levels of CRP and IL-6 were associated with greater odds of prevalent dementia and cognitive impairment non-dementia. Similarly, a case-cohort study (n=727) within the Rotterdam cohort found that increased blood CRP levels were associated with higher risk of dementia (41). Also consistent with our findings, a longitudinal study with a representative sample of 3,075 African American and White participants found that elevated CRP was associated with increased odds of cognitive decline over a two year follow-up period (42). A meta-analysis of pro-inflammatory biomarkers and cognition in older adults found that high levels of IL-6 were associated with cognitive decline, but this study did not find the a similar association with high CRP (43). In contrast, high circulating levels of the anti-inflammatory cytokine IGF-1 are often associated with greater brain volume and lower risk of dementia (44). Indeed, in the Framingham Heart Study (n=3,582), greater IGF-1 levels were associated with lower risk of incident dementia (45). Our cross-sectional results are consistent with these studies, and further suggest that DNA methylation age acceleration may be a biological mechanism linking inflammatory cytokine levels with cognitive impairment.

Our results thus supported our hypothesis that accelerated DNA methylation aging is a plausible mechanism through which inflammatory cytokines influence cognitive status in older adults. DNA methylation clocks have previously been associated with age-related diseases and cognitive decline (14,15,17,46). Accelerated DNA methylation aging may thus be an important mechanism implicated in progressive cognitive deterioration. Our study demonstrated that not only GrimAge acceleration was associated with worsen cognitive status, but that GrimAge acceleration was an intermediate pathway through which CRP, IL-6, and IGF-1 influence cognitive decline. In addition to the GrimAge clock, the DunedinPoAm clock showed an association with dementia, and to a lesser extend with cognitive impairment non-dementia. However, these associations did not meet the threshold of FDR statistical significance. These two phenotypic clocks (GrimAge and DunedinPoAm) have been found to be more predictive of adverse health outcomes such as cognitive decline than the traditional chronological clocks (15,27,46). For example, GrimAge integrates DNA methylation-based estimates of plasma protein levels, including several markers directly associated with smoking pack-years, a significant factor in chronic inflammation and disease risk, which providing a more comprehensive indicator of biological aging compared to other DNA methylation clocks (15,27). Even though the interplay between cytokine exposure and DNA methylation age acceleration is not well documented, mounting evidence underscores the role of age-associated DNA methylation changes as a cellular mechanism linking systemic inflammation to chronic and auto-immune diseases (47,48). In fact, age-related changes in the innate and adaptative immune response are characterized by an overexpression of senescent T cell profiles in circulating blood, the overexpression of the T CD8+ phenotype is a highly regulated epigenetic process and may be induced by cytokines such as CRP and IL-6 (49,50). A recent study exploring trajectories of inflammatory biomarkers, immune cell profiles, and epigenetic ageing in the Lothian Birth Cohort (n=1,091) demonstrated a cross-sectional association between high levels of each of CRP and IL-6 and age-accelerated Hannum residuals (50). Our larger sample size provides enhanced power to detect cross-sectional associations between cytokines and both phenotypic and chronological aging measures compared to these previous studies.

Our analysis is limited by its cross-sectional design. Although, our directed acyclic graph implies a temporal relationship between exposure (cytokine levels), mediator (DNA methylation age acceleration), and outcome (cognitive impairment), these variables were collected at the same time, and hence we are unable to rule out reverse causality. Future research should explore longitudinal trajectories of inflammatory biomarkers and their impact on DNA methylation age acceleration, and cognitive decline. Additionally, we were not able to explore the potential for race-dependent relationships between exposure to cytokines, DNA methylation age acceleration, and adverse cognition, because this study was limited to a subsample of the HRS with a small proportion of participants racialized as non-Hispanic Black or Hispanic. Our limited sample size hindered our ability to conduct stratify our analyses. Our analysis also has several strengths, including a diverse sample of older adults, a large sample size that was greater than most previous studies on this topic, and rich data on well-known confounding variables. Importantly, the HRS has provided cell type proportions, an important confounder given that DNA methylation levels are influenced by the heterogeneity and the cell composition of the tissue in which they are measured, which we were able to use in our statistical models. We also included *APOE-ε4*, a well-known genetic risk factor for dementia. Future studies with time ordered components could shed light on the role of chronic conditions in the inflammatory response of ageing adults and their contribution to cognitive impairment.

In conclusion, our study demonstrated a novel finding on the link between the inflammatory response, cellular processes associated with accelerated aging, and cognitive impairment among older adults. Through mediation analysis, we found that the effect of cytokines in cognitive impairment may be partially mediated through DNA methylation age acceleration. The interplay between persistent inflammatory levels and molecular mechanisms related to premature cognitive aging should further be explored. Future prospective and longitudinal studies should devote attention to understanding the temporal association between these physiological mechanisms to identify what other pathways influence cognitive impairment beyond the dominant amyloid hypothesis.

## Data Availability

All data produced in the present study are available upon reasonable request to the authors.

## Data Availability

Survey data are publicly available, and genetic data are available through dbGaP (https://dbgap.ncbi.nlm.nih.gov; phs000428.v2.p2) or NIAGADS (niagads.org; NG00119 and NG00132. The data presented in this study were accessed from the http://hrs.isr.umich.edu/data-products data repository, Health Data: 2016 Biomarker Data release date November 2020 (Early v1.0).

## Acknowledgments

We thank the participants and staff of the Health and Retirement Study.

## Author Contributions

In alignment with the ICMJE criteria, all authors have made substantial contributions to the conception or design of the work; or the acquisition, analysis, or interpretation of data for the work; AND drafting the work or reviewing it critically for important intellectual content; AND final approval of the version to be published; AND agreement to be accountable for all aspects of the work in ensuring that questions related to the accuracy or integrity of any part of the work are appropriately investigated and resolved.

