## Supplemental Tables and Figures for "DNA Methylation Age Acceleration Mediates the Relationship between Systemic Inflammation and Cognitive Impairment"

This document contains 4 Supplemental Figures, and 4 Supplemental Table.

**Supplemental Figure 1.** Sample inclusion and exclusion flow chart for the Health and Retirement Study 2016 wave.

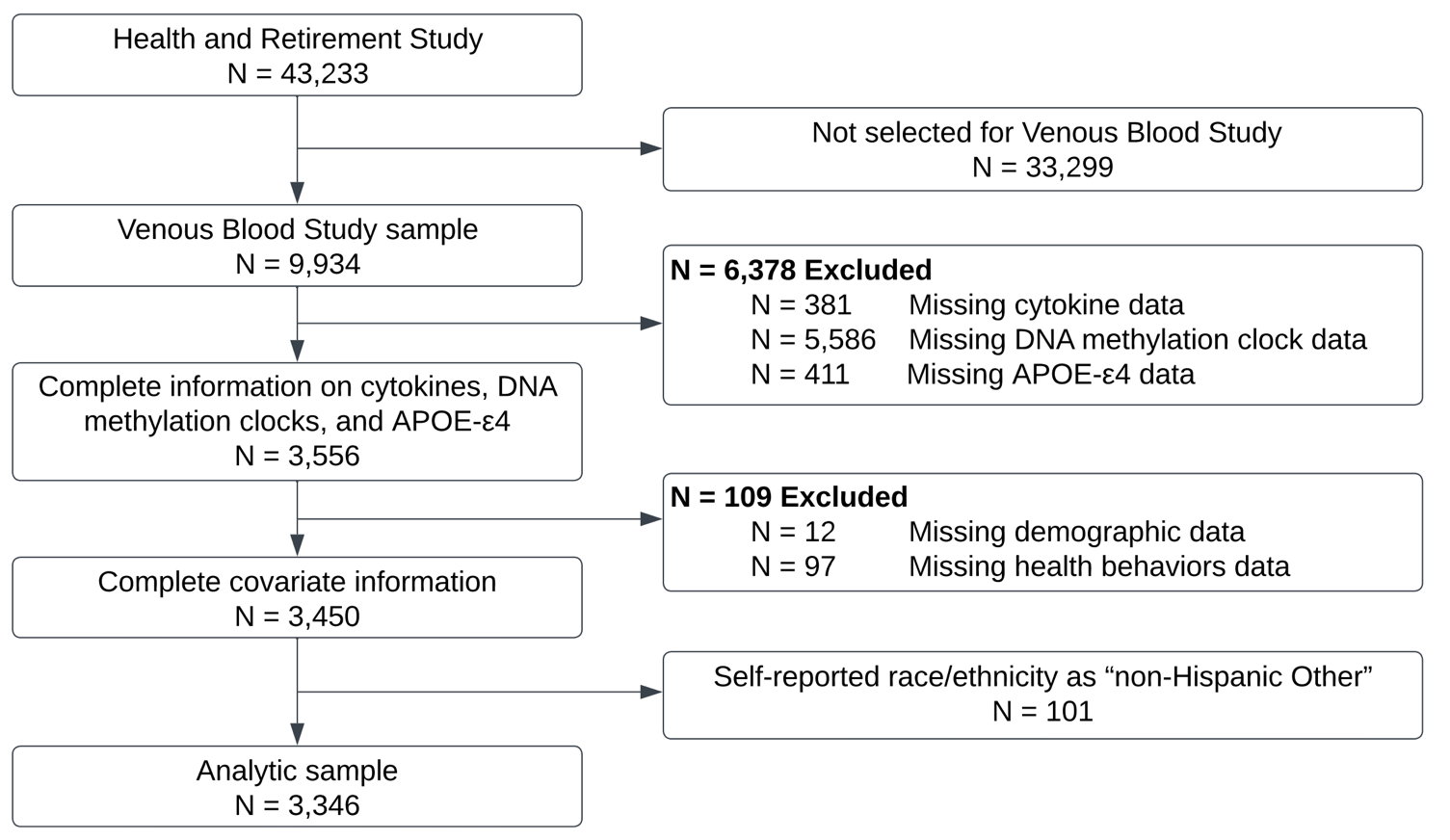

**Supplemental Figure 2:** Correlation between inflammatory cytokine measures (IGF-1, TGF-β1, IL-6, CRP, IL-1RA, IL-10, sTNF-R1) and DNA methylation age acceleration (DunedinPoAm, GrimAge, Levine, Hannum, HorvathSkin, Horvathin the 2016 wave of the Health and Retirement Study. Color (blue to red) reflects the Pearson correlation coefficient.

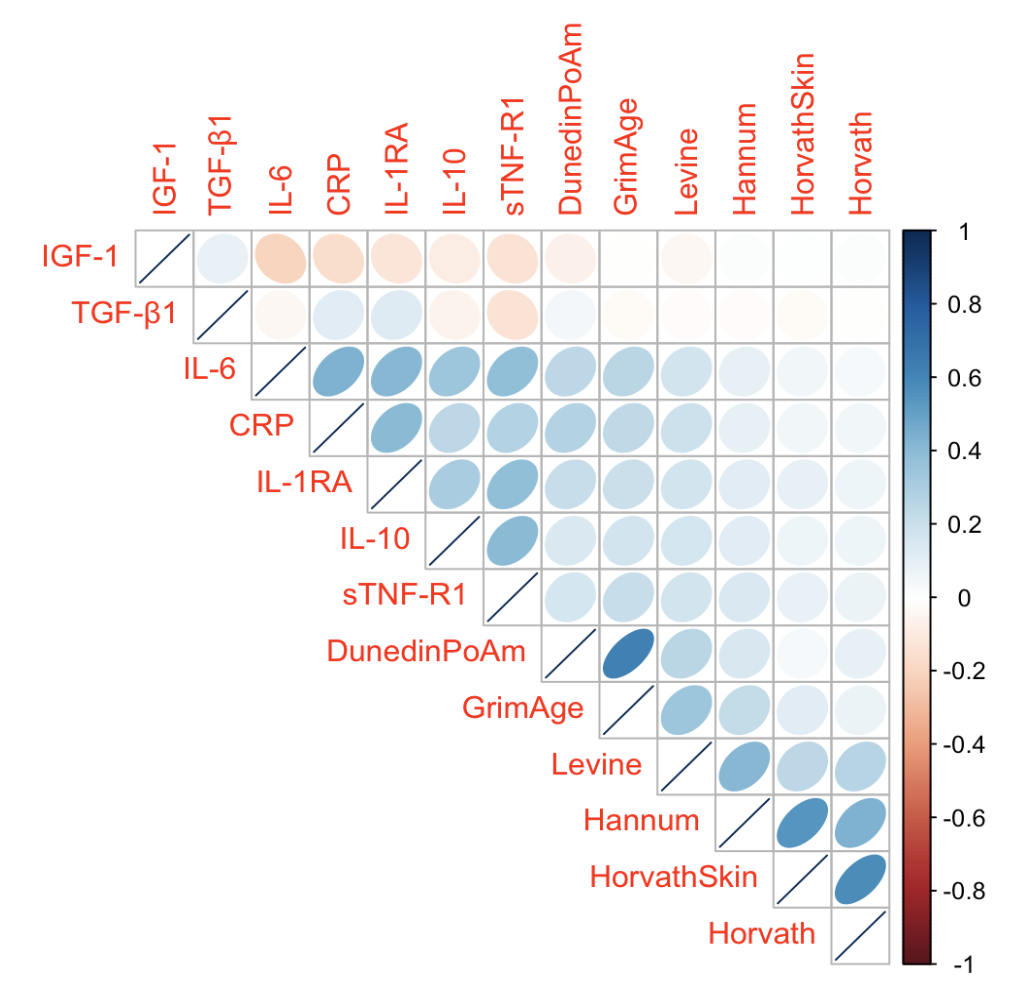

**Supplemental Figure 3:** Forest plot representing the adjusted associations and 95% confidence intervals between cytokine measures and DNA methylation clocks. Beta coefficients were estimated from linear regression analysis and adjusted for sociodemographic characteristics (age, sex, education, race/ethnicity) and cell type proportions (Granulocytes, Monocytes, Lymphocytes) in the 2016 wave of the Health and Retirement Study.

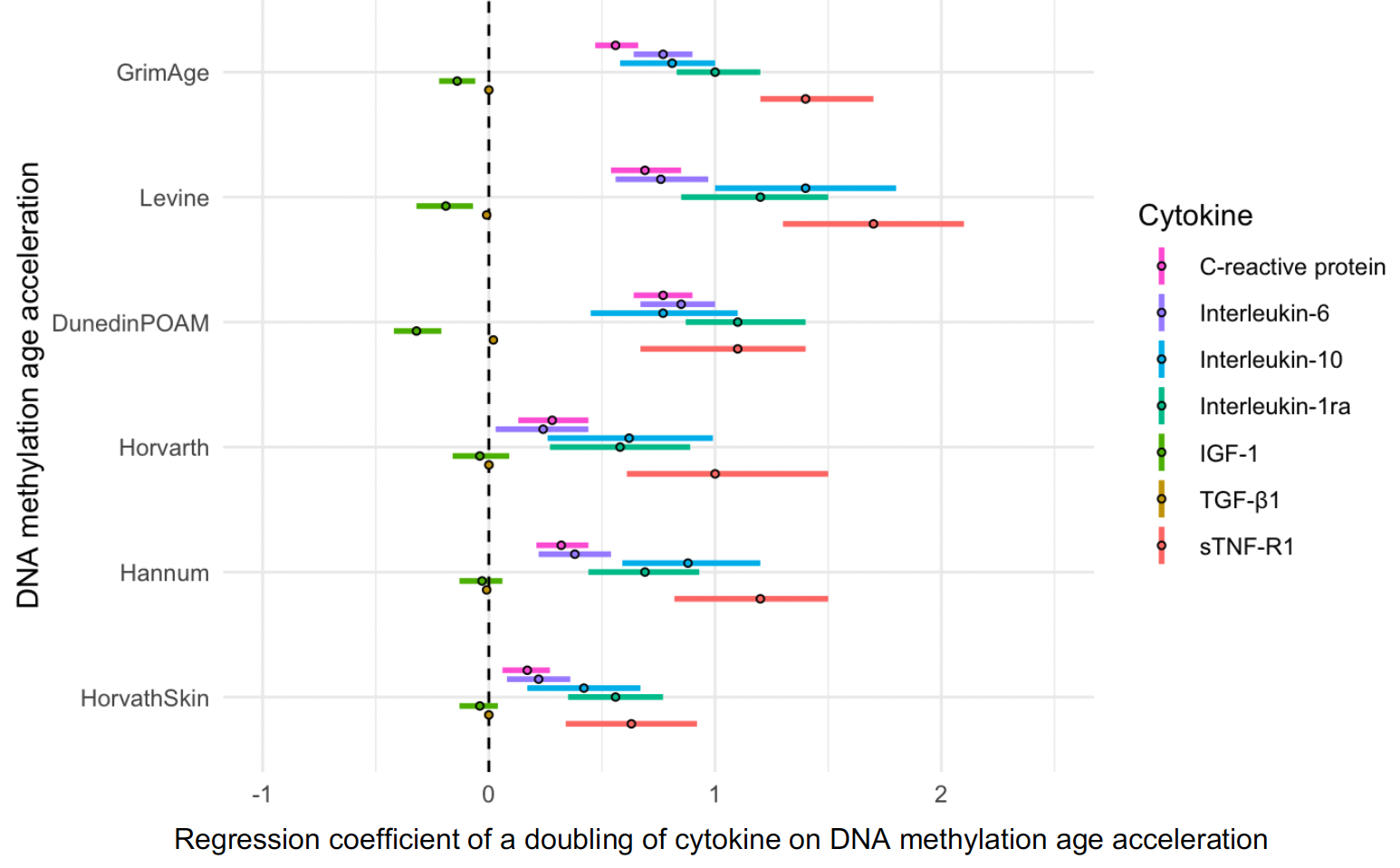

**Supplemental Figure 4:** Scatter plots of chronological age vs. GrimAge and chronological age vs. DNAm GrimAge. Chronological age (years) is plotted on the x-axis. GrimAge clocks and DNAm age (years) predicted by GrimAge clocks are plotted on the y-axis.

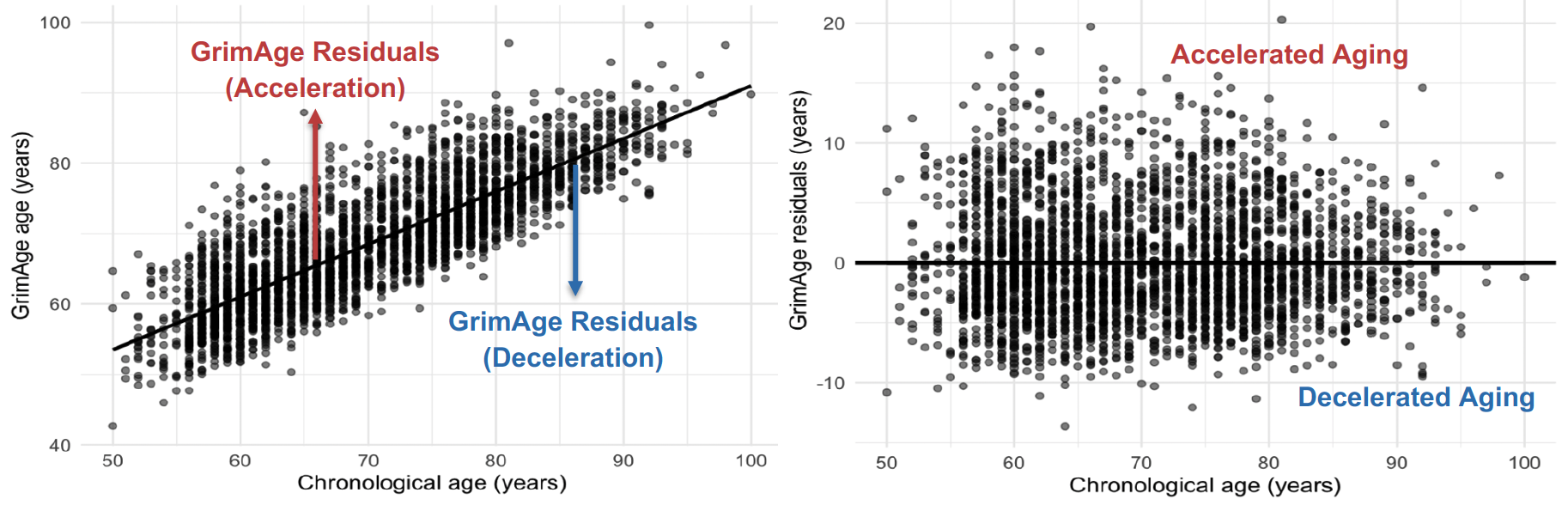

**Supplemental Table 1.** Distribution of participant characteristics among the included and excluded samples of the Health and Retirement Study 2016 wave.

| **Characteristic** | **Excluded Participants**  n = 6,588 | **Analytic Sample Participants**  n = 3,346 |
| --- | --- | --- |
| **Age, years** | 67.81 (10.46) | 69.96 (9.63) |
| **Race/ethnicity** |  |  |
| Non-Hispanic Black | 1,194 (18%) | 554 (17%) |
| Non-Hispanic White | 4,039 (61%) | 2,340 (70%) |
| Other non-Hispanic | 314(4.8%) | 0 |
| Hispanic | 1,024 (16%) | 452 (14%) |
| Unknown | 17 | 0 |
| **Sex** |  |  |
| Female | 3,911 (59%) | 1,957 (58%) |
| Male | 2,677 (41%) | 1,389 (42%) |
| **Educational Category** |  |  |
| > College | 639 (9.7%) | 312 (9.3%) |
| College/Some | 1,467 (22%) | 707 (21%) |
| High school or < | 4,483 (68%) | 2,327 (70%) |
| **Body Mass Index** (kg/m^2^) | 29.14 (6.31) | 28.86 (6.23) |
| Unknown | 110 | 0 |
| **Smoking status** |  |  |
| Current Smoker | 791 (12%) | 361 (11%) |
| Former Smoker | 2,835 (43%) | 1,505 (45%) |
| Never Smoker | 2,909 (45%) | 1,480 (44%) |
| Unknown | 53 | 0 |
| **Exercise** (> 1 time per week) |  |  |
| Yes | 4,571 (70%) | 2,378 (71%) |
| No | 1,979 (30%) | 968 (29%) |
| Unknown | 38 | 0 |
| **Number of drinks per day** | 0.83 (1.48) | 0.81 (1.43) |
| Unknown | 27 | 0 |
| **Number of Chronic Conditions** |  |  |
| ≥ 3 | 2,943 (38%) | 1,510 (45%) |
| 1-2 | 2,910 (44%) | 1,523 (46%) |
| None | 734 (11%) | 313 (9.4%) |
| Unknown | 1 | 0 |

*Notes:* Data presented as either mean (standard deviation) or count (%) where applicable.

**Supplemental Table 2:** Association between inflammatory cytokines and DNA methylation age acceleration in the 2016 venous blood sample of the Health and Retirement Study (n=3,346).

|  | **Phenotypic Clocks** | | | | | | | | | | | **Chronological Clocks** | | | | | | | | | | |
| --- | --- | --- | --- | --- | --- | --- | --- | --- | --- | --- | --- | --- | --- | --- | --- | --- | --- | --- | --- | --- | --- | --- |
| **Cytokine** | **GrimAge** | | | | **Levine** | | | | | **DunedinPoAm** | | **Horvath** | | | | **Hannum** | | | | **HorvathSkin** | | |
|  | **β** | **95%CI** | **FDR** | **β** | | **95%CI** | **FDR** | **β** | **95%CI** | | **FDR** | **β** | **95%CI** | **FDR** | **β** | | **95%CI** | **FDR** | **β** | | **95%CI** | **FDR** |
| **C-reactive protein**^*^ |  |  |  |  | |  |  |  |  | |  |  |  |  |  | |  |  |  | |  |  |
| Model 1 | 0.73 | 0.63, 0.83 | <0.001 | 0.90 | | 0.75, 1.00 | <0.001 | 1.20 | 1.00, 1.30 | | <0.001 | 0.24 | 0.09, 0.38 | 0.002 | 0.32 | | 0.20, 0.44 | <0.001 | 0.15 | | 0.05, 0.25 | 0.004 |
| Model 2 | 0.73 | 0.63, 0.83 | <0.001 | 0.91 | | 0.76, 1.11 | <0.001 | 1.10 | 0.98, 1.30 | | <0.001 | 0.28 | 0.13, 0.43 | <0.001 | 0.42 | | 0.30, 0.53 | <0.001 | 0.18 | | 0.08, 0.28 | <0.001 |
| Model 3 | 0.56 | 0.47, 0.66 | <0.001 | 0.69 | | 0.54, 0.85 | <0.001 | 0.77 | 0.64, 0.90 | | <0.001 | 0.28 | 0.13, 0.44 | <0.001 | 0.32 | | 0.21, 0.44 | <0.001 | 0.17 | | 0.06, 0.27 | 0.002 |
| **Interleukin-6**^*^ |  |  |  |  | |  |  |  |  | |  |  |  |  |  | |  |  |  | |  |  |
| Model 1 | 1.10 | 0.93, 1.20 | <0.001 | 1.00 | | 0.84, 1.20 | <0.001 | 1.40 | 1.20, 1.60 | | <0.001 | 0.22 | 0.03, 0.42 | 0.035 | 0.43 | | 0.27, 0.59 | <0.001 | 0.23 | | 0.10, 0.36 | 0.001 |
| Model 2 | 0.98 | 0.85, 1.10 | <0.001 | 1.10 | | 0.84, 1.30 | <0.001 | 1.30 | 1.10, 1.50 | | <0.001 | 0.24 | 0.04, 0.44 | 0.025 | 0.50 | | 0.34, 0,66 | <0.001 | 0.24 | | 0.10, 0.37 | <0.001 |
| Model 3 | 0.77 | 0.64, 0.90 | <0.001 | 0.76 | | 0.56, 0.97 | <0.001 | 0.85 | 0.67, 1.00 | | <0.001 | 0.24 | 0.03, 0.44 | 0.032 | 0.38 | | 0.22, 0.54 | <0.001 | 0.22 | | 0.08, 0.36 | 0.003 |
| **Interleukin-10**^*^ |  |  |  |  | |  |  |  |  | |  |  |  |  |  | |  |  |  | |  |  |
| Model 1 | 1.40 | 1.10, 1.60 | <0.001 | 1.80 | | 1.40, 2.20 | <0.001 | 1.50 | 1.20, 1.90 | | <0.001 | 0.65 | 0.30, 1.00 | <0.001 | 1.00 | | 0.74, 1.30 | <0.001 | 0.49 | | 0.24, 0.73 | <0.001 |
| Model 2 | 1.10 | 0.87, 1.30 | <0.001 | 1.80 | | 1.40, 2.10 | <0.001 | 1.40 | 1.00, 1.70 | | <0.001 | 0.63 | 0.27, 0.99 | 0.001 | 1.00 | | 0.74, 1.30 | <0.001 | 0.45 | | 0.21, 0.70 | <0.001 |
| Model 3 | 0.81 | 0.58, 1.00 | <0.001 | 1.40 | | 1.00, 1.80 | <0.001 | 0.77 | 0.45, 1.10 | | <0.001 | 0.62 | 0.26, 0.99 | 0.001 | 0.88 | | 0.59, 1.20 | <0.001 | 0.42 | | 0.17, 0.67 | 0.002 |
| **Interleukin-1ra**^*^ |  |  |  |  | |  |  |  |  | |  |  |  |  |  | |  |  |  | |  |  |
| Model 1 | 1.30 | 1.10, 1.50 | <0.001 | 1.60 | | 1.30, 1.90 | <0.001 | 1.90 | 1.60, 2.20 | | <0.001 | 0.54 | 0.24, 0.84 | <0.001 | 0.80 | | 0.56, 1.00 | <0.001 | 0.55 | | 0.34, 0.75 | <0.001 |
| Model 2 | 1.40 | 1.20, 1.60 | <0.001 | 1.60 | | 1.30, 1.90 | <0.001 | 1.90 | 1.60, 2.20 | | <0.001 | 0.57 | 0.27, 0.88 | <0.001 | 0.88 | | 0.64, 1.10 | <0.001 | 0.58 | | 0.37, 0.78 | <0.001 |
| Model 3 | 1.00 | 0.83, 1.20 | <0.001 | 1.20 | | 0.85, 1.50 | <0.001 | 1.10 | 0.87, 1.40 | | <0.001 | 0.58 | 0.27, 0.89 | <0.001 | 0.69 | | 0.44, 0.93 | <0.001 | 0.56 | | 0.35, 0.77 | <0.001 |
| **IGF-1**^†^ |  |  |  |  | |  |  |  |  | |  |  |  |  |  | |  |  |  | |  |  |
| Model 1 | -0.01 | -0.09, 0.08 | 0.9 | -0.13 | | -0.25, -0.01 | 0.047 | -0.22 | -0.33, -0.10 | | <0.001 | 0.05 | -0.07, 0.16 | 0.467 | 0.04 | | -0.06, 0.13 | 0.5 | 0.00 | | -0.08, 0.08 | 0.939 |
| Model 2 | -0.12 | -0.21, -0.04 | 0.004 | -0.17 | | -0.29, -0.04 | 0.013 | -0.28 | -0.40, -0.16 | | <0.001 | -0.03 | -0.16, 0.09 | 0.7 | -0.02 | | -0.12, 0.07 | 0.6 | -0.04 | | -0.12, 0.04 | 0.467 |
| Model 3 | -0.14 | -0.22, -0.06 | <0.001 | -0.19 | | -0.32, -0.07 | 0.004 | -0.32 | -0.42, -0.21 | | <0.001 | -0.04 | -0.16, 0.09 | 0.7 | -0.03 | | -0.13, 0.06 | 0.5 | -0.04 | | -0.13, 0.04 | 0.35 |
| **TGF-β1** |  |  |  |  | |  |  |  |  | |  |  |  |  |  | |  |  |  | |  |  |
| Model 1 | -0.01 | -0.02, 0.00 | 0.23 | -0.01 | | -0.02, 0.01 | 0.5 | 0.02 | 0.00, 0.03 | | 0.011 | 0.00 | -0.02, 0.02 | 0.991 | -0.01 | | -0.02, 0.01 | 0.35 | -0.01 | | -0.02, 0.00 | 0.233 |
| Model 2 | 0.00 | -0.01, 0.01 | 0.991 | 0.00 | | -0.02, 0.01 | 0.6 | 0.02 | 0.01, 0.04 | | 0.002 | 0.00 | -0.01, 0.02 | 0.7 | 0.00 | | -0.02, 0.01 | 0.6 | 0.00 | | -0.01, 0.01 | 0.5 |
| Model 3 | 0.00 | -0.01, 0.01 | 0.5 | -0.01 | | -0.02, 0.01 | 0.3 | 0.02 | 0.00, 0.03 | | 0.015 | 0.00 | -0.01, 0.02 | 0.7 | -0.01 | | -0.02, 0.01 | 0.467 | 0.00 | | -0.02, 0.01 | 0.4 |
| **sTNF-R1**^*^ |  |  |  |  | |  |  |  |  | |  |  |  |  |  | |  |  |  | |  |  |
| Model 1 | 1.80 | 1.60, 2.10 | <0.001 | 2.10 | | 1.70, 2.50 | <0.001 | 1.90 | 1.50, 2.30 | | <0.001 | 0.91 | 0.53, 1.30 | <0.001 | 1.30 | | 0.99, 1.60 | <0.001 | 0.64 | | 0.37, 0.90 | <0.001 |
| Model 2 | 1.90 | 1.60, 2.20 | <0.001 | 2.30 | | 1.90, 2.70 | <0.001 | 2.10 | 1.70, 2.50 | | <0.001 | 1.00 | 0.60, 1.40 | <0.001 | 1.40 | | 1.10, 1.70 | <0.001 | 0.66 | | 0.38, 0.95 | <0.001 |
| Model 3 | 1.40 | 1.20, 1.70 | <0.001 | 1.70 | | 1.30, 2.10 | <0.001 | 1.10 | 0.67, 1.40 | | <0.001 | 1.00 | 0.61, 1.50 | <0.001 | 1.20 | | 0.82, 1.50 | <0.001 | 0.63 | | 0.34, 0.92 | <0.001 |

*Notes:* CI = confidence interval in brackets; FDR = false discovery rate. IGF-1 = Insulin-Like Growth Factor 1; sTNF-R = soluble Tumoral Necrosis Factor; TGF-ß1 = Transforming Growth Factor Beta 1. Model 1: Unadjusted; Model 2: adjusted for race/ethnicity, age, sex, education; Model 3: adjusted for race/ethnicity, age, sex, education categories, and cell type proportions (percent granulocytes and lymphocytes) coefficient estimated from linear regression.

^*^ Variable log base 2 transformed.

^†^ Squared root transformed.

**Supplemental Table 3:** Regression-based mediation analysis estimates for cytokine exposures associated with cognitive impairment as the outcome using GrimAge acceleration as mediator. Models are presented for each cytokine separately in a sample of the Health and Retirement Study (N=3,346).

|  | **C-reactive protein** | | | **Interleukin-6** | | | **Insulin-Like Growth Factor-1** | | |
| --- | --- | --- | --- | --- | --- | --- | --- | --- | --- |
| **Rate Ratio Scale** | **Estimate** | **Lower bound** | **Upper bound** | **Estimate** | **Lower bound** | **Upper bound** | **Estimate** | **Lower bound** | **Upper bound** |
| Controlled Direct Effect | 1.06 | 1.00 | 1.14 | 1.09 | 1.02 | 1.18 | 0.94 | 0.90 | 0.98 |
| Pure Natural Direct Effect | 1.06 | 1.00 | 1.14 | 1.09 | 1.02 | 1.18 | 0.94 | 0.91 | 0.98 |
| Total Natural Direct Effect | 1.06 | 1.00 | 1.14 | 1.09 | 1.02 | 1.18 | 0.94 | 0.91 | 0.98 |
| Pure Natural Indirect Effect | 1.02 | 1.01 | 1.02 | 1.02 | 1.01 | 1.03 | 1.00 | 0.99 | 1.00 |
| Total Natural Indirect Effect | 1.02 | 1.01 | 1.02 | 1.02 | 1.01 | 1.03 | 1.00 | 0.99 | 1.00 |
| Total Effect | 1.07 | 1.01 | 1.15 | 1.11 | 1.04 | 1.20 | 0.94 | 0.90 | 0.98 |
| Portion Mediated | 0.23 | 0.04 | 0.97 | 0.18 | 0.07 | 0.51 | 0.06 | 0.01 | 0.19 |
| **Mediational E-value** | **Estimate** | **Lower bound** | **Upper bound** | **Estimate** | **Lower bound** | **Upper bound** | **Estimate** | **Lower bound** | **Upper bound** |
| Pure Natural Indirect Effect | 1.14 | NA | 1.08 | 1.15 | NA | 1.09 | 1.07 | 1.03 | NA |

*Notes:* Outcome is cognitive impairment (dementia and cognitive impairment non-dementia combined). Models adjusting for socio-demographic variables (age, sex, racialized social group, education), behavioral factors (body mass index, exercise, smoking status, alcohol consumption), chronic conditions, APOE-ε4 allele carrier status, and cell type proportions (percent granulocytes and lymphocytes).

**Supplemental Table 4:** Prevalent Odds Ratios (ORs) from logistic regression analysis, representing the direct effect association for each doubling of cytokine levels (CRP, IL-6, IGF-1) with prevalent cognitive impairment, relative to normal cognition in the US Health and Retirement Study (n=3,346).

|  | **Cognitive impairment (non-dementia and dementia) vs. Normal cognitive function** | | | | | |
| --- | --- | --- | --- | --- | --- | --- |
|  | **C-reactive protein**^*^ | | **Interleukin-6**^*^ | | **Insulin-Like Growth Factor-1**^†^ | |
| **Variables** | **OR** | **[95%CI]** | **OR** | **[95%CI]** | **OR** | **[95%CI]** |
| **C-reactive protein**^*^ | 1.07 | 1.00, 1.14 | - | - | - | - |
| **Interleukin-6**^*^ | - | - | 1.11 | 1.01, 1.21 | - | - |
| **Insulin-Like Growth Factor-1**^†^ | - | - | - | - | 0.93 | 0.88, 0.98 |
| **GrimAge** | 1.04 | 1.02, 1.07 | 1.04 | 1.02, 1.07 | 1.05 | 1.02, 1.08 |
| **Race** |  |  |  |  |  |  |
| Non-Hispanic White | - | - | - | - | - | - |
| Non-Hispanic Black | 2.93 | 2.25, 3.81 | 2.91 | 2.24, 3.79 | 3.05 | 2.34, 3.97 |
| Hispanic | 3.41 | 2.61, 4.46 | 3.39 | 2.59, 4.43 | 3.28 | 2.50, 4.29 |
| **Gender** |  |  |  |  |  |  |
| Female | - | - | - | - | - | - |
| Male | 1.31 | 1.06, 1.62 | 1.30 | 1.05, 1.60 | 1.32 | 1.07, 1.64 |
| **Age** | 1.07 | 1.05, 1.08 | 1.06 | 1.05, 1.08 | 1.06 | 1.05, 1.08 |
| **Education** |  |  |  |  |  |  |
| > College | - | - | - | - | - | - |
| College / some college | 3.42 | 1.82, 7.04 | 3.47 | 1.85, 7.14 | 3.47 | 1.85, 7.12 |
| High school or less | 7.07 | 3.94, 14.1 | 7.19 | 4.01, 14.3 | 6.99 | 3.90, 13.9 |
| **APOE-ε4 allele status** |  |  |  |  |  |  |
| No Copy | - | - | - | - | - | - |
| At least 1 copy | 1.54 | 1.24, 1.90 | 1.50 | 1.21, 1.84 | 1.50 | 1.22, 1.85 |
| **Granulocytes** | 1.01 | 0.97, 1.05 | 1.01 | 0.97, 1.05 | 1.01 | 0.97, 1.05 |
| **Lymphocytes** | 1.01 | 0.97, 1.05 | 1.01 | 0.97, 1.05 | 1.01 | 0.97, 1.05 |
| **BMI** | 0.96 | 0.94, 0.97 | 0.96 | 0.94, 0.98 | 0.96 | 0.94, 0.98 |
| **Exercise** |  |  |  |  |  |  |
| Yes | - | - | - | - | - | - |
| No | 1.44 | 1.18, 1.76 | 1.44 | 1.18, 1.76 | 1.46 | 1.20, 1.79 |
| **Smoking status** |  |  |  |  |  |  |
| Never | - | - | - | - | - | - |
| Current smoker | 0.84 | 0.57, 1.23 | 0.83 | 0.56, 1.21 | 0.82 | 0.56, 1.21 |
| Former smoker | 0.92 | 0.74, 1.14 | 0.92 | 0.74, 1.14 | 0.93 | 0.75, 1.15 |
| **Alcohol consumption / day** | 0.82 | 0.75, 0.89 | 0.82 | 0.75, 0.90 | 0.82 | 0.75, 0.89 |
| **Chronic conditions** |  |  |  |  |  |  |
| None | - | - | - | - | - | - |
| 1-2 | 1.00 | 0.68, 1.50 | 1.02 | 0.69, 1.52 | 1.01 | 0.69, 1.52 |
| >=3 | 1.29 | 0.87, 1.93 | 1.29 | 0.88, 1.94 | 1.31 | 0.89, 1.97 |

*Notes:* OR = Odds Ratio; CI = confidence interval in brackets; FDR = false discovery rate.

^*^ Variable log base 2 transformed.

^†^ Squared root transformed.
